# Enhancing MRI Safety: Real-Time Thermal Imaging Integrated with Deep Learning for Burn Prevention

**DOI:** 10.1101/2025.11.09.25339848

**Authors:** Selah McKenney, Edward Walsh, James Webb, Jeffery Rogg, Michael Oumano

## Abstract

**Background:** Radiofrequency (RF)-induced burns are the most common MRI-related adverse event. Standard safety practices such as visual checks and patient communication are often insufficient, especially for anesthetized or incapacitated patients.

**Purpose:** To evaluate the feasibility of combining thermal infrared imaging with a convolutional neural network (CNN) for detecting abnormal heating in real time during MRI.

**Methods:** Phantom and human subject experiments were performed on 3T MRI systems. A thermal camera captured images from front and back scanner placements during controlled heating. Images were labeled, preprocessed, and used to train CNNs in MATLAB. Model performance was assessed by accuracy, sensitivity, specificity, and area under the curve (AUC).

**Results:** Phantom testing demonstrated linear temperature rise with constant specific absorption rate (SAR) scanning. In human subject testing, CNNs achieved accuracies between 78–100%. Front placement yielded higher performance, likely due to larger datasets and improved visibility. A radar plot summarizing network performance demonstrated robust classification.

**Conclusions:** This study demonstrates the feasibility of CNN-enhanced infrared monitoring to detect abnormal heating during MRI, providing a potential path toward automated, real-time burn prevention systems.

## Introduction

### 1 The Evolution and Risks of Magnetic Resonance Imaging

Magnetic resonance imaging (MRI) is an indispensable diagnostic tool, offering unparalleled soft-tissue contrast without ionizing radiation. Since the 1970s, MRI technology has become central to modern clinical practice, particularly for imaging of neurological, cardiovascular, and oncologic disease (1,2).

Despite its advantages, MRI is associated with safety risks, most notably RF-induced burns. These result from conductive loops, proximity effects, or interactions with devices. RF heating increases with the square of frequency, and thus with scanner field strength (3).

Patient burns are the most commonly reported adverse events in MRI, accounting for ∼59% of all MRI-related incidents in FDA records (4). These burns are largely preventable, yet continue to occur despite established guidelines. One reason is that safety strategies such as patient communication and technologist visual checks fail when patients are anesthetized, pediatric, or unable to report discomfort.

Previous work has explored infrared (IR) thermography as a quality assurance tool for coils (5), but no studies have implemented automated thermal monitoring of patients in real time.

The FDA’s MAUDE database documents more than 100 RF burns in the last three years, with many cases involving coil cables, bore contact, and monitoring loops (7). Approximately three-quarters of these burns would likely have been visible to an infrared thermal camera, suggesting a clear opportunity for proactive monitoring.

This study presents a proof-of-concept system combining IR imaging with a CNN to detect hotspots during MRI. The aim was to evaluate feasibility, quantify detection performance, and identify implementation pathways for clinical use.

## Methods

### 2.1 Thermal Camera Performance Index and Quality Assurance

The infrared thermal camera used in this study was the Kaiweets KTI-W01, equipped with a vanadium oxide uncooled focal plane array. It offered a resolution of 256 × 192 pixels, with a pixel size of 12 µm and an infrared response range of 8 to 14 µm. The camera featured a 3.2 mm focal length lens and a thermal measurement resolution of 0.1 °C.

Quality assurance (QA) testing was performed to ensure MRI compatibility (Figure 1). A MAGNETOM Skyra Fit 3T scanner (Siemens Healthineers) equipped with a 32-channel head coil (BioMatrix) was used. A standard MRI phantom containing all 32 coils was placed on the patient couch during these tests.

**Figure 1.**
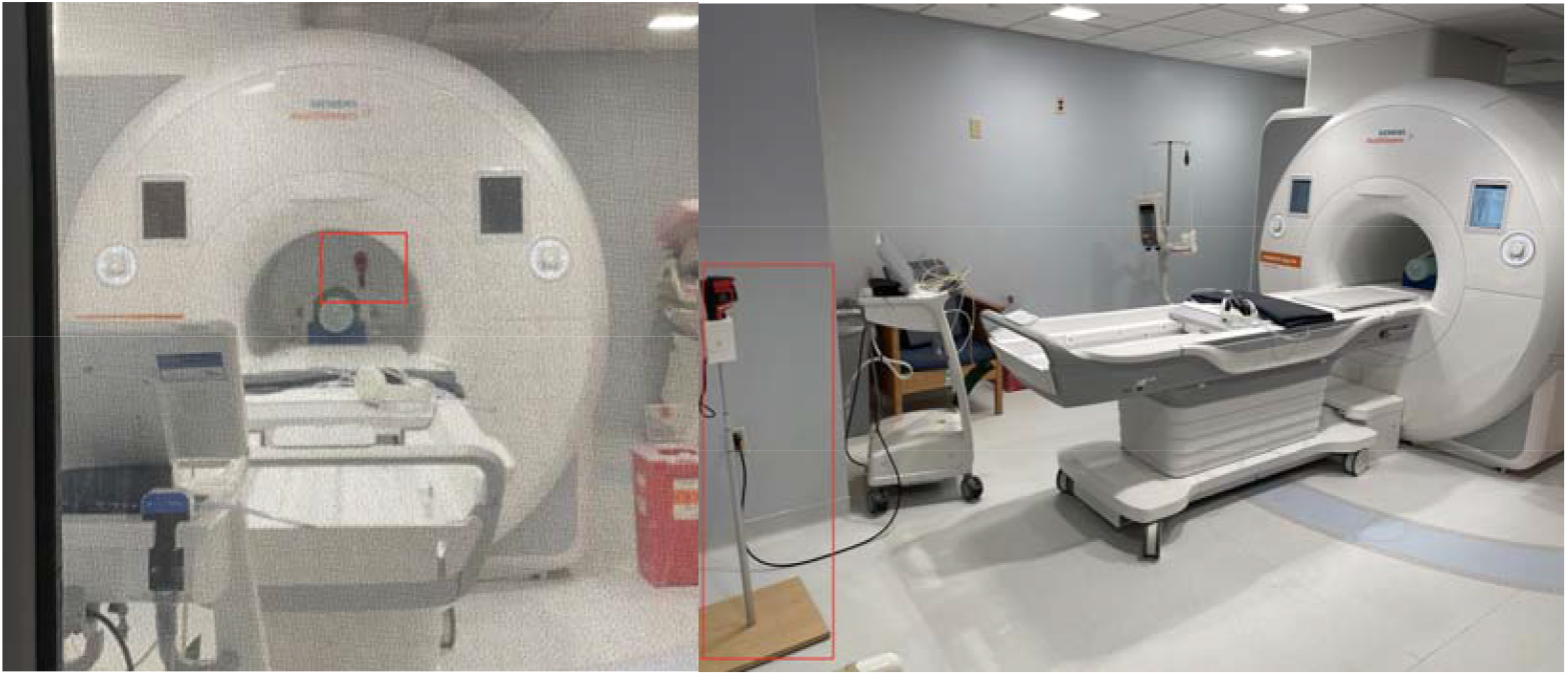
“Back” and “Front” Camera Placement. Back Placement (left image): The camera was positioned behind the scanner, outside the 400 Gauss line. Front Placement (right image): The camera was positioned in front of the scanner, outside the 5 Gauss line.

The tests were conducted under four different conditions:

**1. Baseline QA:** Performed without the camera or stand in the room.

**2. Front (Camera Off):** The camera and stand were positioned in front of the scanner, but the camera remained powered off.

**3. Front (Camera On):** The camera and stand remained in the same position, but the camera was powered on.

**4. Back (Camera On):** The camera remained powered on but was repositioned to the back of the scanner.

These tests confirmed that the presence and operation of the thermal imaging system did not interfere with the MRI system’s performance.

Each coil on the head phantom was evaluated for signal-to-noise ratio (SNR), brightness, and transmitter amplitude. Each coil had specific tolerance ranges for these values. Across all tested camera positions, including the baseline, front (camera off), front (camera on), and back (camera on), the recorded values remained within acceptable tolerances. The following graphs (Supplemental 1A-1C) demonstrate that the addition of the camera did not significantly affect any of the QA test parameters for individual coils, confirming that the infrared imaging system did not interfere with MRI performance.

### 2.2 Heat Pack in Contact with Bore

To simulate a potential patient burn scenario, a commercial heat pack (12” x 18”) filled with a mixture of water, a thickening agent, a freezing agent, and silica gel was securely taped (yellow and black striped tape) to the inner wall of the MRI bore (Figures 2-3)[6]. This setup was designed to mimic inadvertent heating from patient-skin contact with the bore liner. A standard clinical imaging sequence was then run to induce background RF energy deposition and simulate real-use conditions. The MRI protocol used was a transverse T2-weighted turbo spin echo (T2 TSE) sequence with the following parameters: repetition time (TR) = 4420 ms, echo time (TE) = 8.3 ms, 1 average, 1 concatenation, field of view (FOV) = 220 mm × 220 mm, matrix size = 352 × 352, slice thickness = 3 mm, flip angle = 180°, turbo factor = 60, bandwidth = 406 Hz/Px, echo spacing = 8.31 ms, Gradient Mode = Fast, and Specific Absorption Rate (SAR) set to Normal Operating Mode (2 W/kg body limit averaged over 15 minutes). Thermal camera data were recorded throughout the sequence to capture heat propagation.

**Figure 2.**
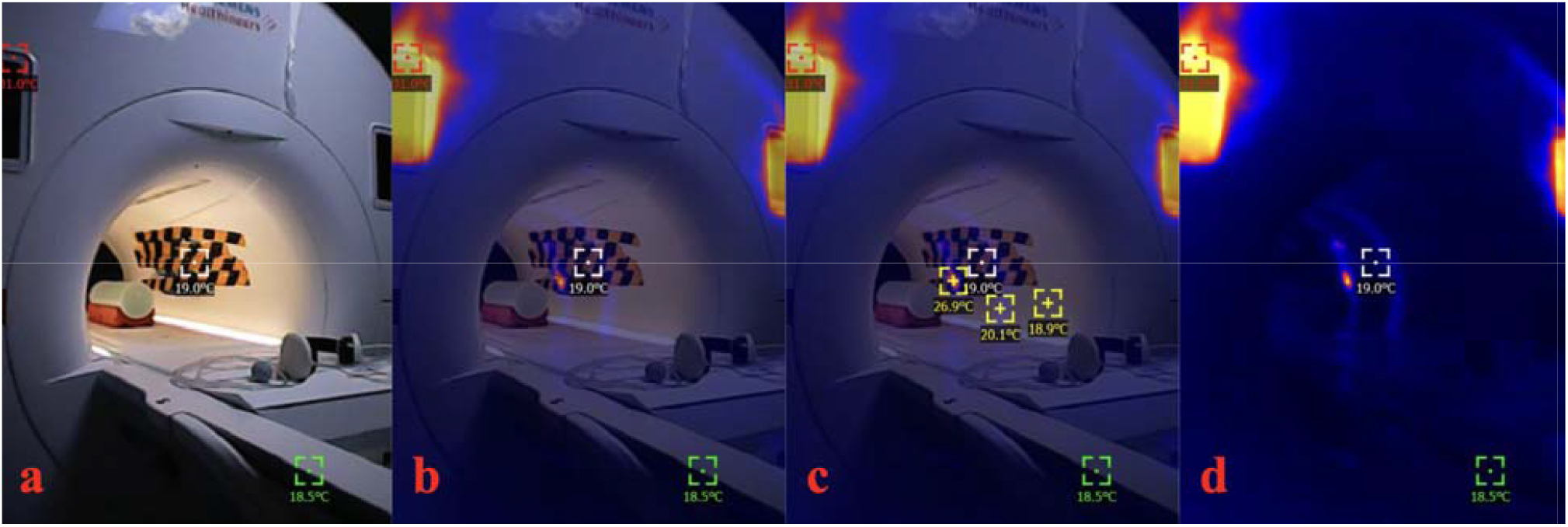
Heat pack in contact with bore and camera positioned at front of the MRI scanner. (a) RGB only (b) RGB-thermal fused (c) RGB-thermal fused with additional temperature signatures (d) Thermal only.

Note the main body transmit coils cause two thermal rings within the bore. The most intense heating occurred when the heat pack was in contact with one of these rings.

Just as in Figure 2 above, there are two thermal rings within the bore. Again, the most intense heating occurred where the heat pack was in contact with one of these rings.

Following the sequence, the heat pack was removed and the thermal camera was used to image the temperature of the heat pack (Figure 4).

**Figure 3.**
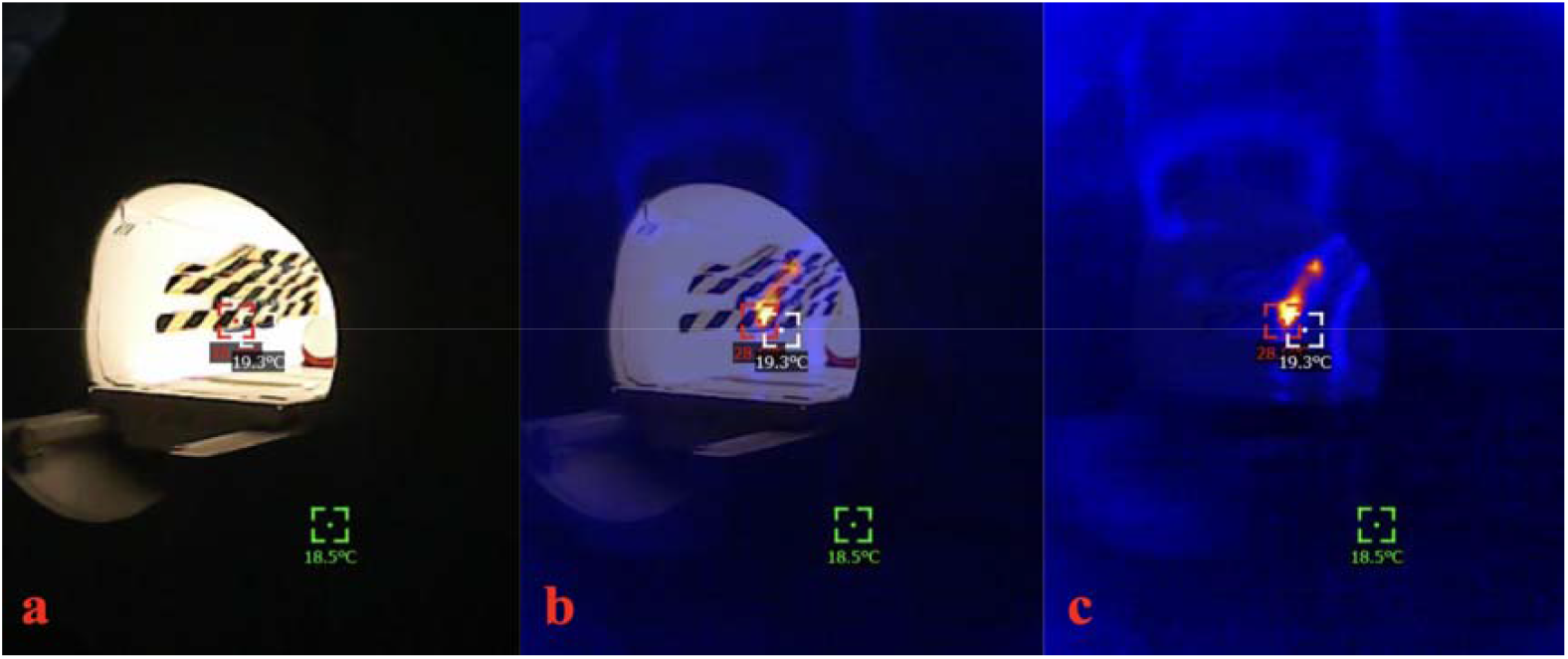
Heat pack in contact with bore and camera positioned at back of the MRI scanner. (a) RGB only (b) RGB-thermal fused (c) Thermal only.

**Figure 4.**
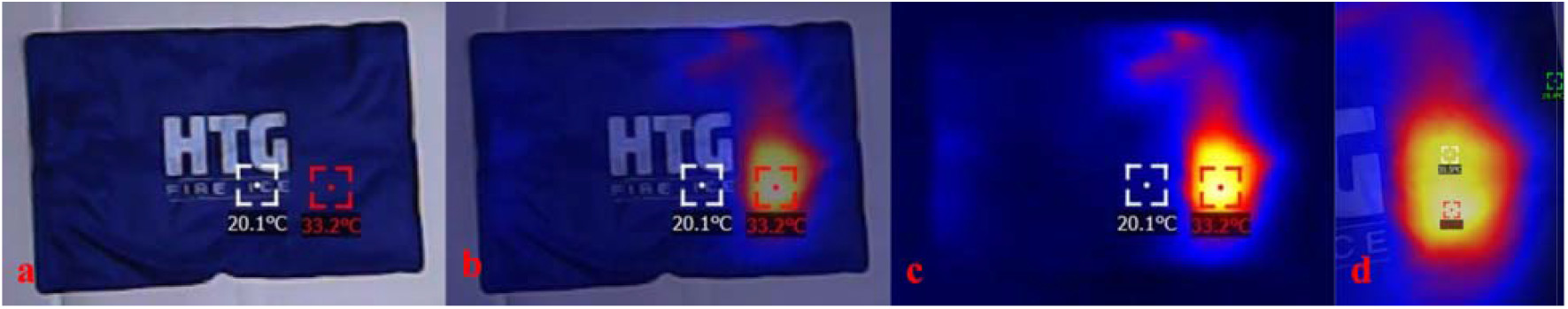
Heat pack after MRI sequence heating. (a) RGB only (b) RBG-thermal fused (c) thermal only (d) zoomed-in image of hot spot in contact with main body transmit coil

### 2.3 Human Subject

A human subject was used to simulate a real-world MRI scan on a General Electric (GE) Signa Premier 3T (Figure 5). Institutional review board (IRB) approval was obtained via IRB protocol number 2262636-2. The subject was positioned supine on the MRI table, with heated vials and heat packs placed at various anatomical locations. To replicate a clinical head MRI scan, a head coil was initially positioned on the table for part of the image acquisition and was subsequently removed.

**Figure 5.**
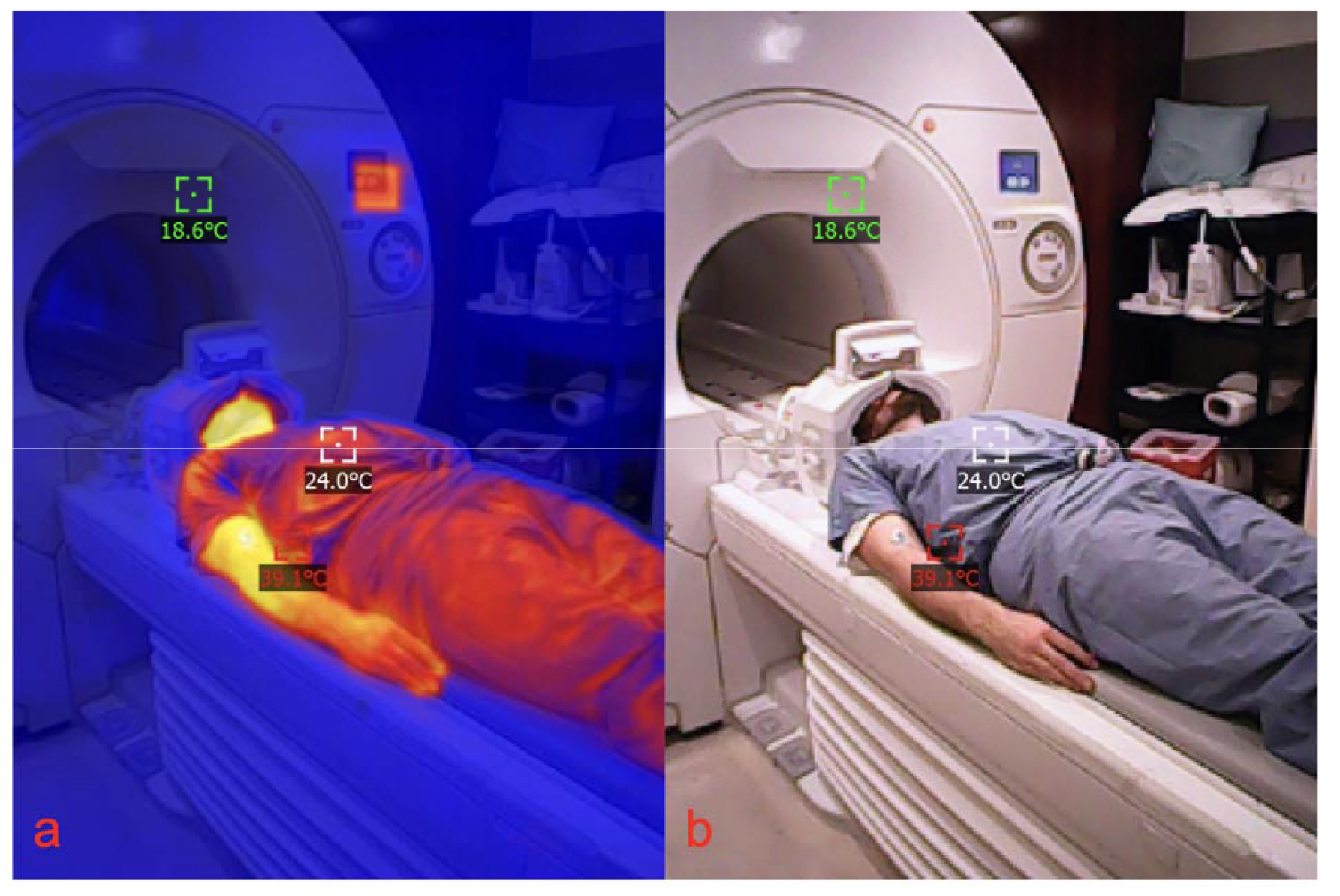
Infrared image of human subject in 3T MRI scanner (a) IR image (b) RBG image

### 2.4 Generation of Unnatural Tissue Heating Images

A standardized protocol was developed to guide the placement of warm vials and heat packs on various anatomical locations for thermal imaging data collection in an MRI scanner. The procedure involves heating the materials to temperatures between 30°C and 55°C—above typical human skin temperature—and systematically capturing images at 20 locations using two couch positions (A and B) from both “front” and “back” perspectives.

This results in approximately 800 images per subject (400 front, 400 back). Images captured with heat sources are classified as ‘hot,’ while those taken without heat sources are categorized as ‘cold’.

### 2.5 Preparation of Heat Sources

Non-conductive 1 mL vials and rectangular heat packs (5 × 3 inches) are heated in a water bath maintained at approximately 60°C (140°F). Upon removal, the vials exhibit temperatures approximately 10-25°C above typical human surface temperature. After about 5 minutes, they are returned to the water bath, and a new vial is selected for use. As shown in Figure 6, after about 10 minutes, the vials become thermally indistinguishable from human skin temperature.

**Figure 6.**
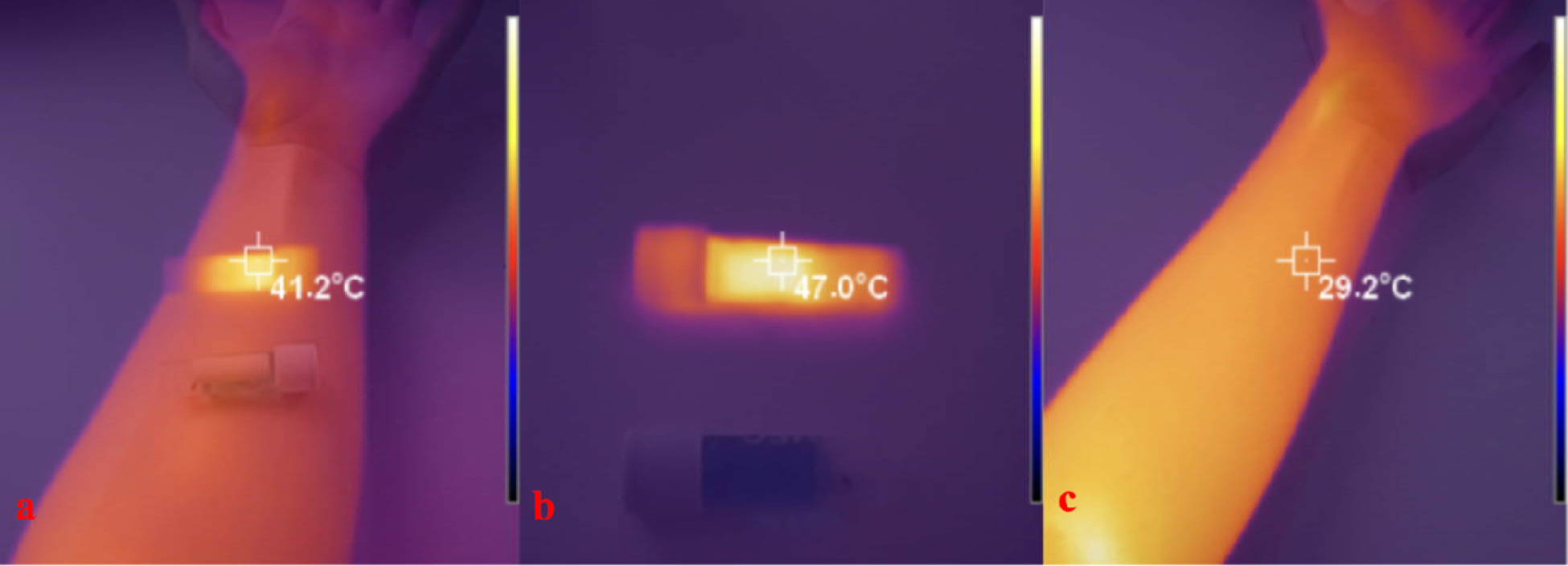
Infrared images of a heated vial (a) heated vial against human skin (b) heated vial in air (c) heated vial against human skin after 10 minutes.

### 2.6 Application of Heat Sources

Once the heat sources reach the desired temperature, they are immediately placed on the subject’s body in a predefined order. Vials are placed at 12 specific locations, including the forehead, neck, chest (between the pectorals), left and right shoulders, elbows, hands, quadriceps, and feet (Figure 7). Heat packs are applied to eight locations, including the head, neck, chest, left and right elbows, and quadriceps. The placement alternates between Couch Position A and Couch Position B to capture a comprehensive dataset.

**Figure 7.**
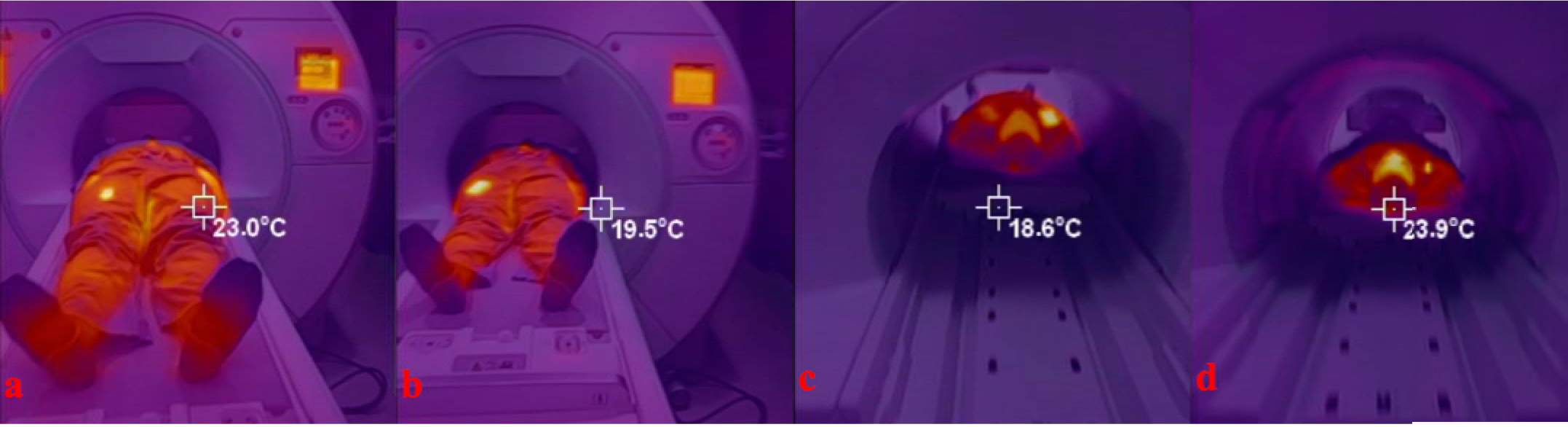
Human subject with vial and pack heat sources examples of “hot” images. (a) Vial heat source on left quadricep at front camera placement at couch position A (b) Pack heat source on left quadricep at front camera placement at couch position A (c) Vial heat source on right shoulder at back camera placement at couch position A (d) Pack heat source on right shoulder at back camera placement at couch position A

### 2.7 Imaging Procedure

Each body site was photographed 10 times per couch position at the front of the MRI scanner, then repeated at the back to maintain alignment consistency. Heat sources (vials and packs heated to 30–55 °C in a water bath) were re-heated every 5 minutes to maintain above-skin temperatures.

### 2.8 Image Selection

Only images with visible heat sources were included for CNN training and testing. For the front camera placement, head and neck sites were sometimes obscured, while for the back placement, lower body regions and those obstructed by the head coil were excluded. This resulted in modest differences in dataset sizes across networks.

### 2.9 Model Development

A CNN was implemented in MATLAB (R2024b, Deep Learning Toolbox) to classify thermal images as “hot” or “cold.” Training optimization included increasing epochs, lowering the base learning rate, and adding dynamic learning rate scheduling to improve convergence while minimizing overfitting. Batch sizes varied with dataset size (typically 32–64 images per batch).

### 2.10 CNN Architecture

Input RGB images (720 × 1280 × 3) were processed through successive convolutional, batch normalization, ReLU, and max-pooling layers. The first layer applied 8 filters (3 × 3 kernel), followed by 16 and 32 filters in subsequent layers. Feature maps were progressively downsampled from 720 × 1280 to 90 × 160. Flattened feature maps were passed into a fully connected layer with two output nodes (hot vs cold) and a softmax classifier. Training used stochastic gradient descent with momentum (SGDM), a learning rate of 0.0005, L2 regularization of 0.0001, and 50 epochs with scheduled halving every 5 epochs.

### 2.11 Human Subject Networks

Six CNNs were trained, divided by camera placement (front or back) and heat source type (vials, packs, or mixed). Equal numbers of training and testing images were used across networks to allow direct comparisons of performance (Table 1).

**Table 1.** Summary of Data for Human Subject CNNs.

| Network Name | Scanner Position | Image Type | Total Images | Training Images | Testing Images |
| --- | --- | --- | --- | --- | --- |
| Human Vial Front | Front | Only vial images | 540 | 420 | 120 |
| Human Pack Front | Front | Only heat pack images | 540 | 420 | 120 |
| Human Front Mix | Front | 50% vials, 50% heat packs | 540 | 420 | 120 |
| Human Vial Back | Back | Only vial images | 400 | 300 | 100 |
| Human Pack Back | Back | Only heat pack images | 400 | 300 | 100 |
| Human Back Mix | Back | 50% vials, 50% heat packs | 400 | 300 | 100 |

## Results

### 3.1 Heat Pack in Contact with Bore

Figure 8 below shows a linear increase in temperature with time. Figure 4 above indicates that the actual final heat pack temperature was closer to 33.5 degrees Celsius, but Figure 3 indicates a final temperature of 28.6 deg C from the back. Considering the additional RF energy deposited during the time spent moving the camera from the front (Figure 2) to the back of the MRI scanner (Figure 3), these results are all in agreement. However, thermal cameras are often more accurate when closer to the heat source, due to pixel averaging at greater distances, which helps explain the 4.9 degree difference between Figure 4d and Figure 3c.

**Figure 8.**
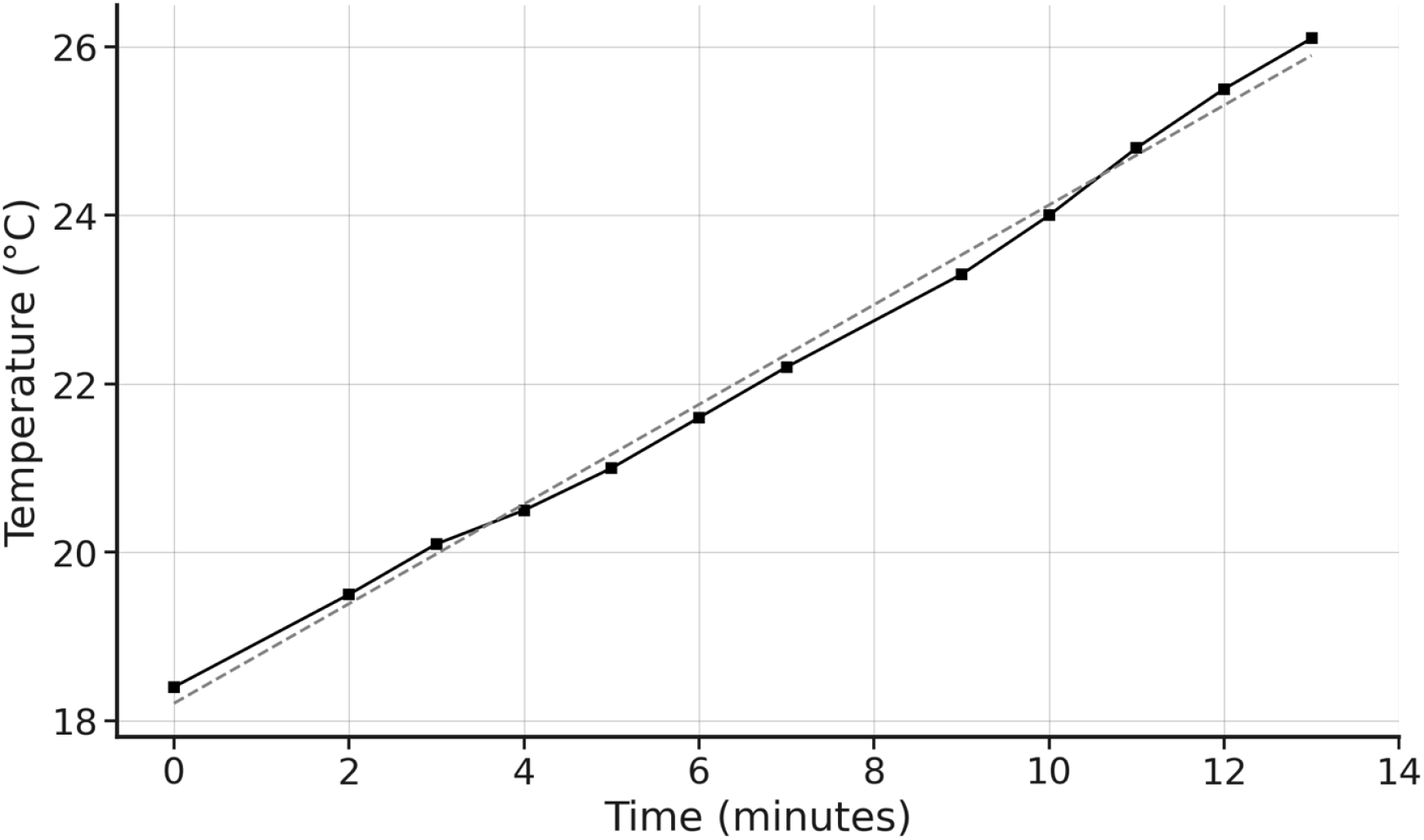
Temperature rise of heat pack during MRI sequence as measured from the front. Linear regression: y = 0.5611x + 18.383, R^2^ = 0.9967. During MRI scanning, the temperature rise is linear over time because the RF power deposition (SAR) is constant, resulting in a steady rate of energy absorption by the tissue or heat pack in this case. In the early phase of heating, before significant heat dissipation through conduction or perfusion occurs, this steady energy input leads to a proportional, linear increase in temperature.

**Figure 9.**
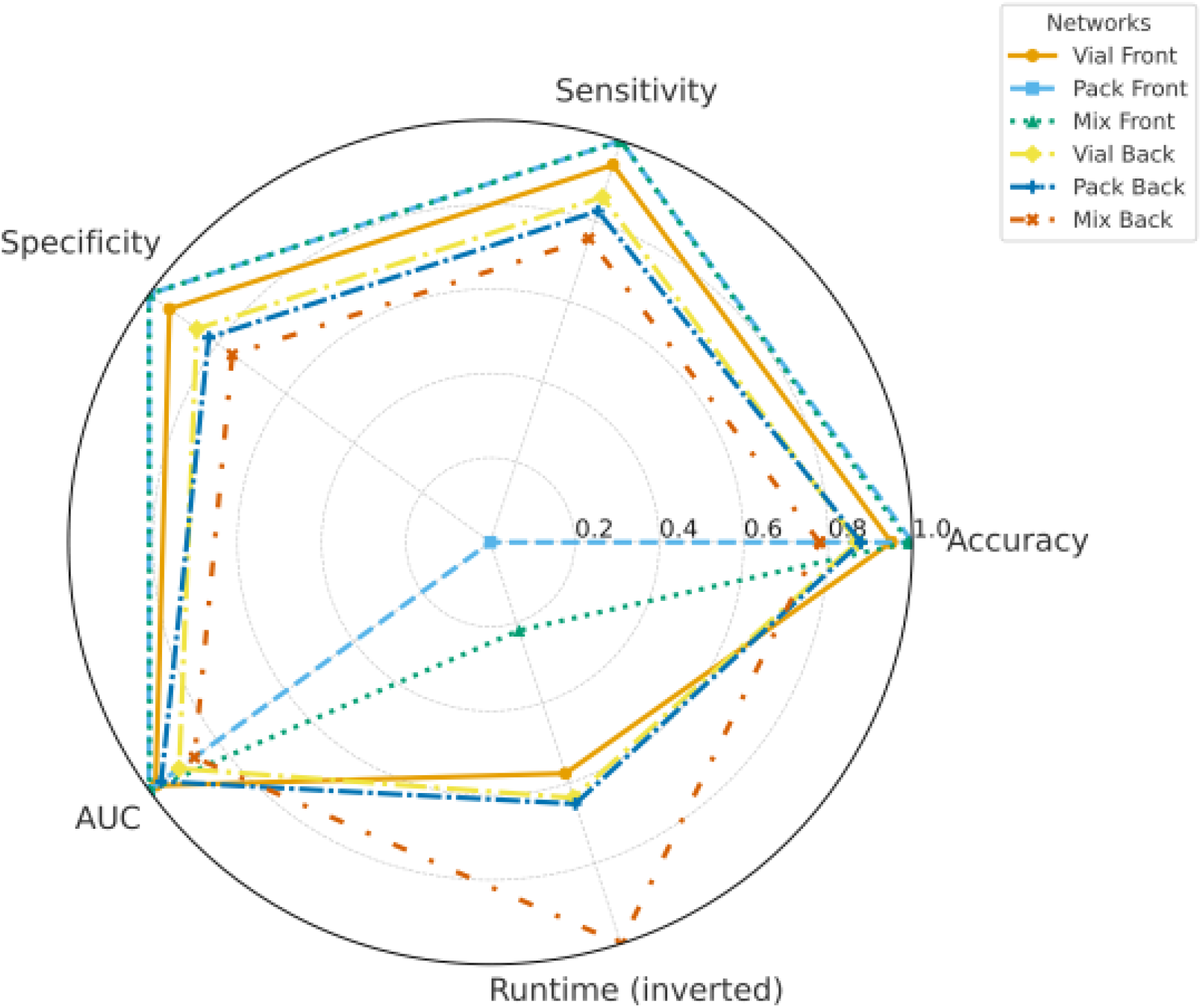
Radar plot summarizing CNN performance across six human subject networks. Metrics shown include accuracy, sensitivity, specificity, AUC, and inverted runtime (higher radial values represent shorter runtime). Each line represents a separate CNN trained on images from different scanner placements and heat source types: Vial Front (solid line with circles), Pack Front (dashed line with squares), Mix Front (dotted line with triangles), Vial Back (dash-dot line with diamonds), Pack Back (long-dash line with pentagons), and Mix Back (spaced-dash line with X markers).

**Figure 10.**
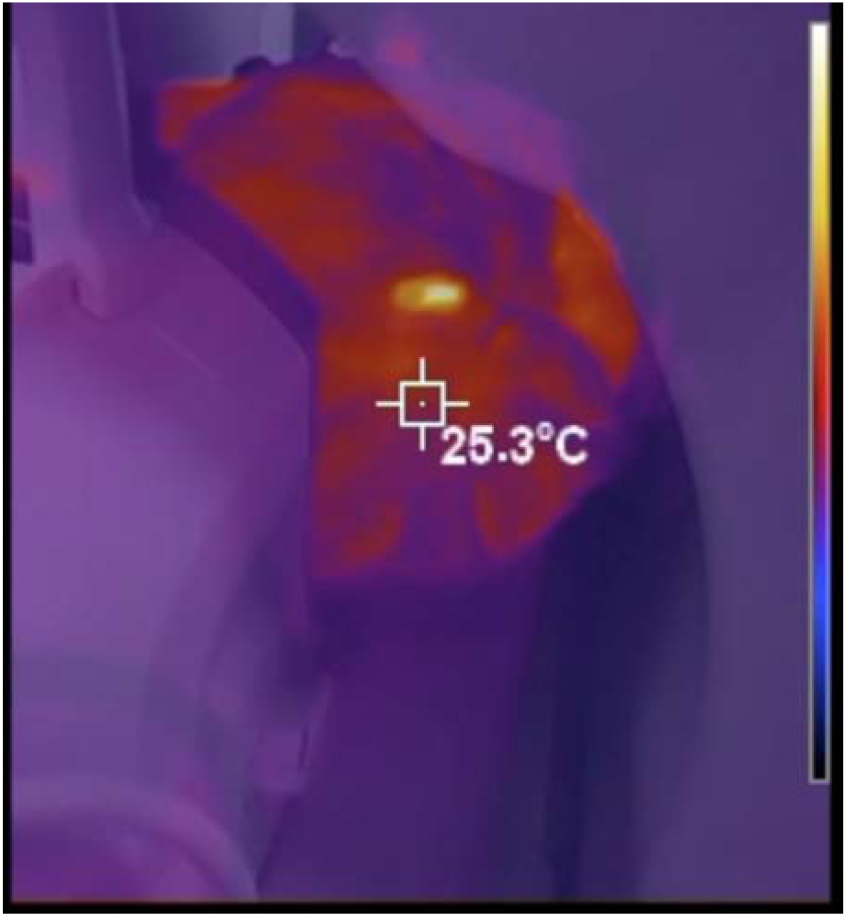
Example of a “hot” image of the human subject with head coil and camera at the back of the MRI scanner.

### 3.2 CNN Detection of Heat Sources on Human Subject

For each CNN trained on the human subject data, multiple statistical tests were conducted to evaluate the performance of binary image classification. These tests assessed the model’s ability to detect the presence of a hot object on the human subject, as well as the time required for training and testing.

As shown in Table 2 below, the front-view networks generally outperformed the back-view networks but required a longer runtime. Additionally, the front-view network underwent more iterations (150) compared to the back-view network (100). The number of iterations represents the batches processed within a single epoch. Although the total number of epochs—the number of times the dataset is passed through the model—remained the same for both networks, the batch size varied due to differences in the number of available images. The larger dataset associated with the front-view networks resulted in a greater batch size, leading to the observed difference in the number of iterations.

**Table 2.** Summary of results for human subject networks.

| Network Name | Run Time | Total Images | Iterations | Epochs | Learning Rate | Accuracy | Sensitivity | Specificity | AUC |
| --- | --- | --- | --- | --- | --- | --- | --- | --- | --- |
| Vial Front | 10:51:58 | 540 | 150 | 50 | 9.77E-07 | 0.95 | 0.94 | 0.94 | 0.9804 |
| Pack Front | 18:27:46 | 540 | 150 | 50 | 9.77E-07 | 1 | 1 | 1 | 1 |
| Mix Front | 15:38:00 | 540 | 150 | 50 | 9.77E-07 | 0.9916 | 1 | 1 | 0.99796 |
| Vial Back | 10:07:38 | 400 | 100 | 50 | 9.77E-07 | 0.87 | 0.86 | 0.86 | 0.9118 |
| Pack Back | 9:51:47 | 400 | 100 | 50 | 9.77E-07 | 0.8778 | 0.825 | 0.825 | 0.9645 |
| Mix Back | 5:19:30 | 400 | 100 | 50 | 9.77E-07 | 0.7802 | 0.7561 | 0.7561 | 0.86683 |

The findings of this study underscore the importance of real-time monitoring to prevent RF-induced burns during MRI. High-power RF fields can induce currents, cause ohmic heating, and elevate tissue temperatures, particularly in areas where skin contacts skin or the bore. Our results show that AI-automated detection using infrared thermography can identify localized hotspots in real time. By integrating a thermal camera with a CNN, the system can flag heating risks and alert technologists, or in future applications, directly interrupt scanning to prevent injury. The model achieved 78–100% accuracy (Figure 4) in detecting hotspots from both front and back camera placements.

### 4.1 Camera Placement

Detection performance was strongly influenced by camera position. The front placement provided a broader field of view and more training data, while back placement was limited to the chest, shoulders, head, and neck. To overcome this, multiple cameras could be used at the front and back of the scanner. Images without visible heat sources were excluded, leading to differences in dataset size and complicating direct accuracy comparisons. Importantly, only anatomy inside the bore is at risk of RF heating; areas outside are not relevant for safety monitoring. Future systems should prioritize coverage of regions exposed to RF deposition.

### 4.2 RF-Induced Heating and Patient Injuries

A review of FDA MAUDE reports identified 102 MRI burns since March 2022, including injuries from cables, monitor loops, ECG leads, transducers, and bore contact. Some cases required surgical intervention. Notably, several burns occurred in anesthetized or sedated patients, where communication-based protocols were ineffective. Of these events, 72.5% would likely have been visible to a thermal camera, particularly bore-contact injuries. However, internal or implanted conductors would not be detectable, a limitation of this technique.

### 4.3 Head Coil

The head coil was present in 29% of “hot” images in the back-camera dataset. Despite partial obstruction, the CNN still detected heating when sources were within view. Misclassifications were unrelated to coil presence, suggesting that the model reliably recognized heat sources above skin temperature. Heat sources obscured from the back view could still be visualized from the front, supporting the use of dual-camera monitoring.

Future research will focus on improving model accuracy, employing higher-resolution thermal cameras, optimizing integration into clinical workflows, and testing across a broader range of MRI conditions and patient scenarios. Ideally, MRI suites would use multiple thermal cameras at the front and back of the scanner. A real-time video feed could be analyzed frame by frame by the CNN, which would flag high-temperature regions and alert technologists, or in future implementations, automatically halt the scan.

### 4.4 Future Directions

#### Temporal Resolution

Future work should improve the system’s ability to detect rapid temperature changes, which may occur over seconds in high-risk scenarios such as wire loops or improperly decoupled coils. Enhancing update rates would enable earlier intervention before burns occur. Dynamic adjustment of update intervals could balance accuracy with computational performance. Physiological factors such as perfusion also affect risk: poorly perfused regions, such as extremities in vascular-compromised patients, may heat faster and should be considered in system design and model training.

#### Camera Placement and Detection

Camera positioning is critical. Infrared-reflective mirrors could extend coverage to areas otherwise obscured, with algorithms correcting for distortion. Deploying multiple cameras at strategic locations (e.g., scanner ends or room corners) could further improve monitoring and hotspot detection.

#### Expanding Testing Protocols

Future studies should include larger and more diverse datasets across body sizes, anatomical sites, and coil configurations. Broader testing will be essential to evaluate robustness and ensure generalization across patient populations.

## Conclusion

Automated thermal monitoring using infrared imaging and CNN classification is a feasible approach for enhancing MRI safety. Phantom and human subject studies demonstrate robust detection of abnormal heating patterns. With further optimization and clinical validation, this system could provide MRI technologists with real-time alerts, reducing the incidence of RF-induced burns and improving patient safety.

## Data Availability

All data produced in the present study are available upon reasonable request to the authors

## Acknowledgments

The authors acknowledge the support of Brown Health for providing access to MRI scanners used in this study. During the preparation of this work the authors used *ChatGPT (OpenAI)* to assist with text editing, improving clarity, and formatting in accordance with journal requirements. After using this tool, the authors reviewed and edited the content as needed and take full responsibility for the content of the published article.

## Declaration of Interest

The authors declare that they have no known competing financial interests or personal relationships that could have appeared to influence the work reported in this paper.

## Figures

Figure 1. Camera placement configurations within the MRI suite.

(A) Camera positioned behind the scanner (“Back”), outside the 400 Gauss line.

(B) Camera positioned in front of the scanner (“Front”), outside the 5 Gauss line. Abbreviations: MRI, magnetic resonance imaging; Gauss, magnetic flux density unit.

Figure 2. Heat pack contact scenario, front camera placement.

Thermal and RGB-thermal fused images show elevated heating where the heat pack touches the inner bore wall, near transmit coils.

Figure 3. Heat pack contact scenario, back camera placement.

Thermal and RGB-thermal images confirm consistent heating patterns corresponding to coil location.

Figure 4. Post-MRI thermal analysis of heat pack.

(a) RGB image; (b) RGB-thermal fused; (c) thermal only; (d) zoomed hot spot detail, confirming a peak surface temperature of 33.5°C.

Figure 5. Human subject in MRI scanner with thermal imaging.

(a) Infrared image; (b) RGB image during simulated scan protocol.

Figure 6. Vial temperature decay over time.

(a) Heated vial on skin; (b) vial in air; (c) vial on skin after 10 min, showing thermal dissipation and convergence with skin temperature.

Figure 7. “Hot” example images from vial and pack placements.

Examples of thermal imaging with varied positioning, camera angles, and heat source types.

Figure 8. Figure 8: Temperature rise of heat pack during MRI sequence as measured from the front. Linear regression: y = 0.5611x + 18.383, R^2^ = 0.9967. During MRI scanning, the temperature rise is linear over time because the RF power deposition (SAR) is constant, resulting in a steady rate of energy absorption by the tissue or heat pack in this case. In the early phase of heating, before significant heat dissipation through conduction or perfusion occurs, this steady energy input leads to a proportional, linear increase in temperature.

Figure 9. Radar plot summarizing CNN performance across six human subject networks. Metrics shown include accuracy, sensitivity, specificity, AUC, and inverted runtime (higher radial values represent shorter runtime). Each line represents a separate CNN trained on images from different scanner placements and heat source types: Vial Front (solid line with circles), Pack Front (dashed line with squares), Mix Front (dotted line with triangles), Vial Back (dash-dot line with diamonds), Pack Back (long-dash line with pentagons), and Mix Back (spaced-dash line with X markers).

Figure 10. Human subject image showing head coil during scan.

Thermal detection accuracy was not significantly impacted by partial visual occlusion from the head coil.

## Supplemental Tables

**Table 1.** MAUDE MRI Burn Reports Summary.

| Source of Burn | # Cases Likely Detectable by IR | % Detectable |
| --- | --- | --- |
| Bore contact | 33 | 100% |
| Coil cables | 21 | 88% |
| ECG leads | 9 | 75% |
| Stimulator interference | 4 | 0% |
| Other | 7 | 60% |
| Total | 74/102 | 72.5% |

## Supplemental Figures

**Figure 1A.**
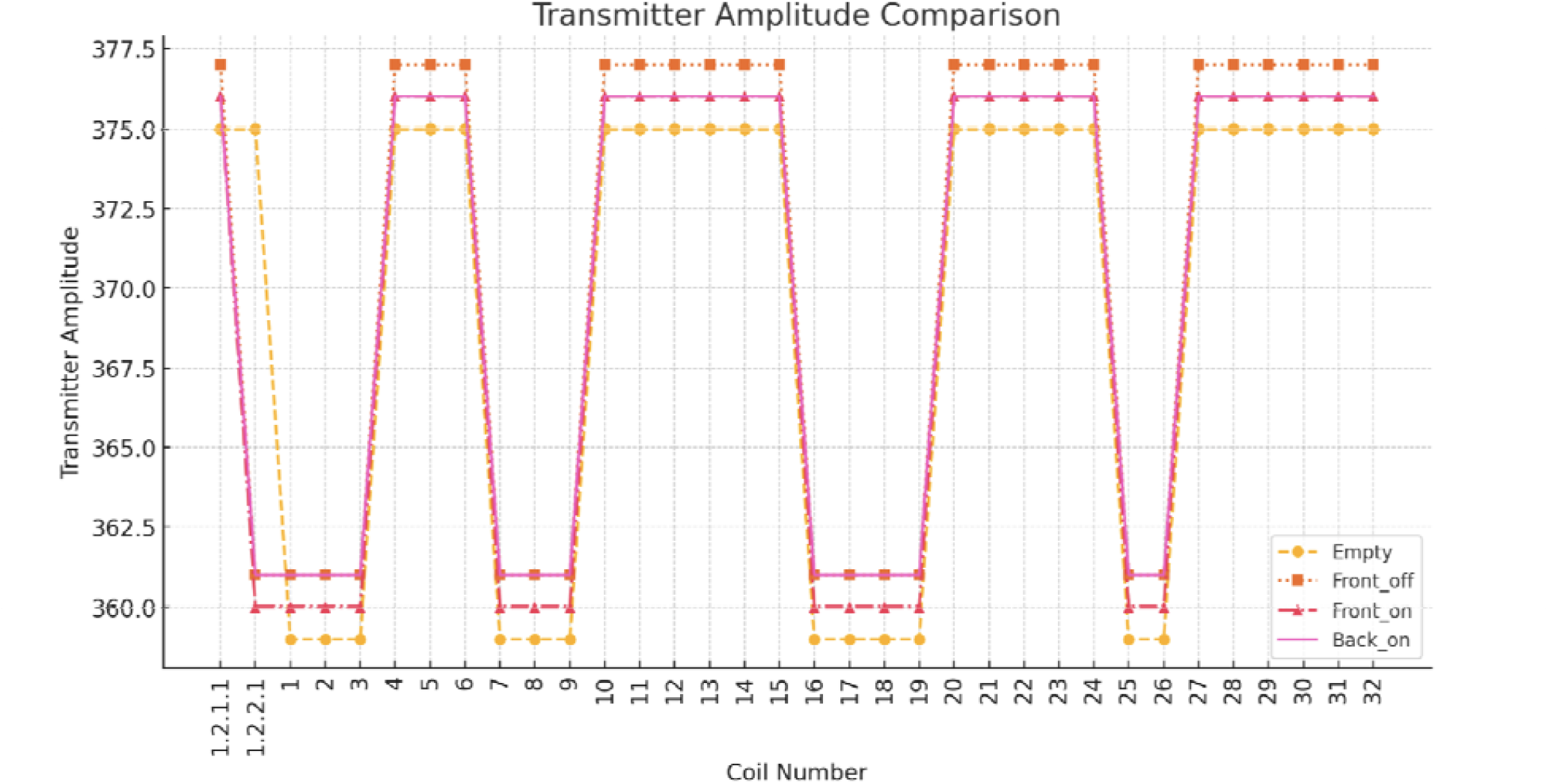
: Quality Assurance of Head Phantom Transmitter Amplitude Comparison conditions: no camera or stand (empty), camera off in front of room (front_off), camera on in front of the room (front_on), camera on in the back of the room (back_on)

**Figure 1B.**
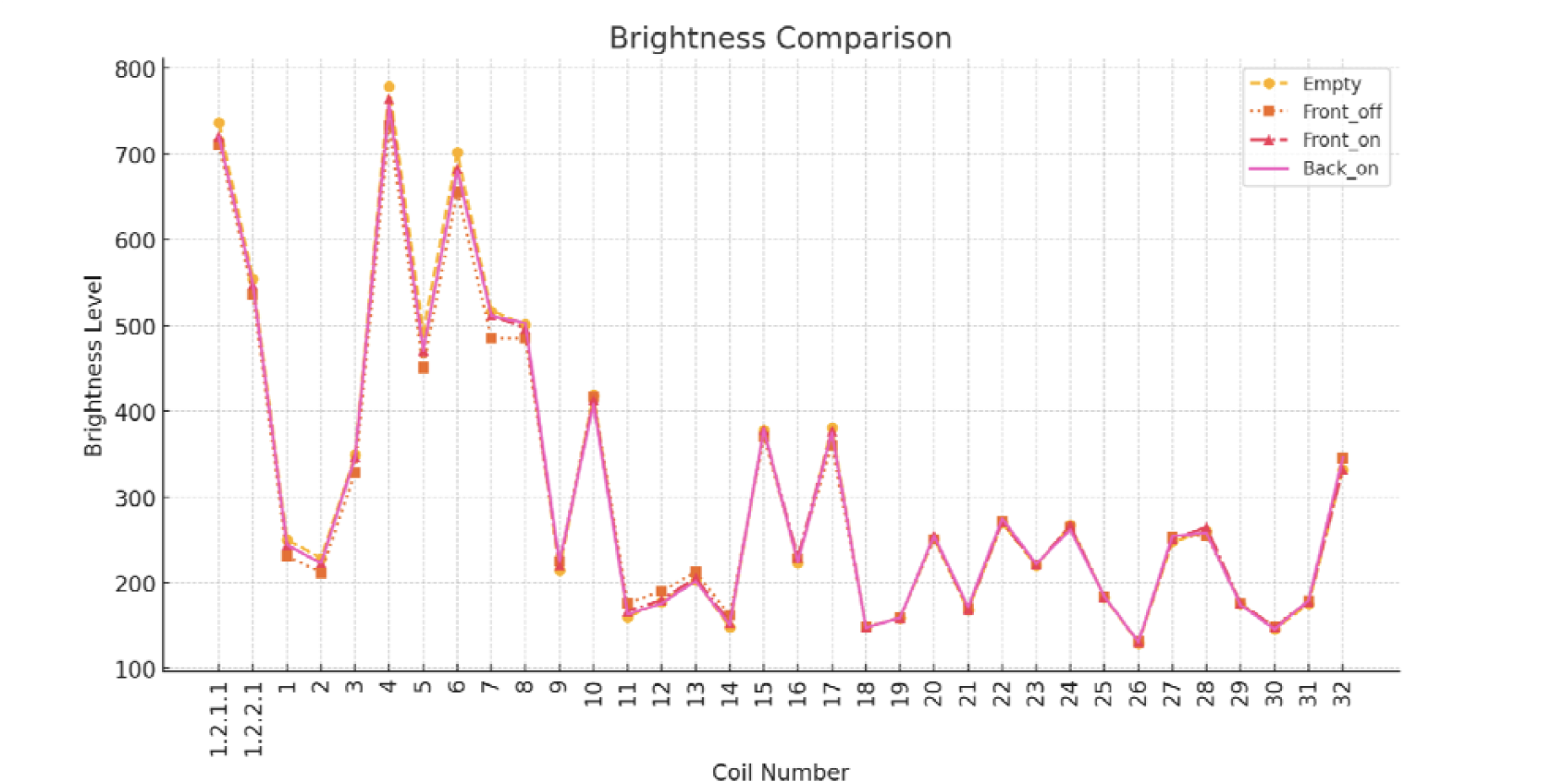
Quality Assurance of Head Phantom Brightness Comparison conditions: no camera or stand (empty), camera off in front of room (front_off), camera on in front of the room (front_on), camera on in the back of the room (back_on)

**Figure 1C.**
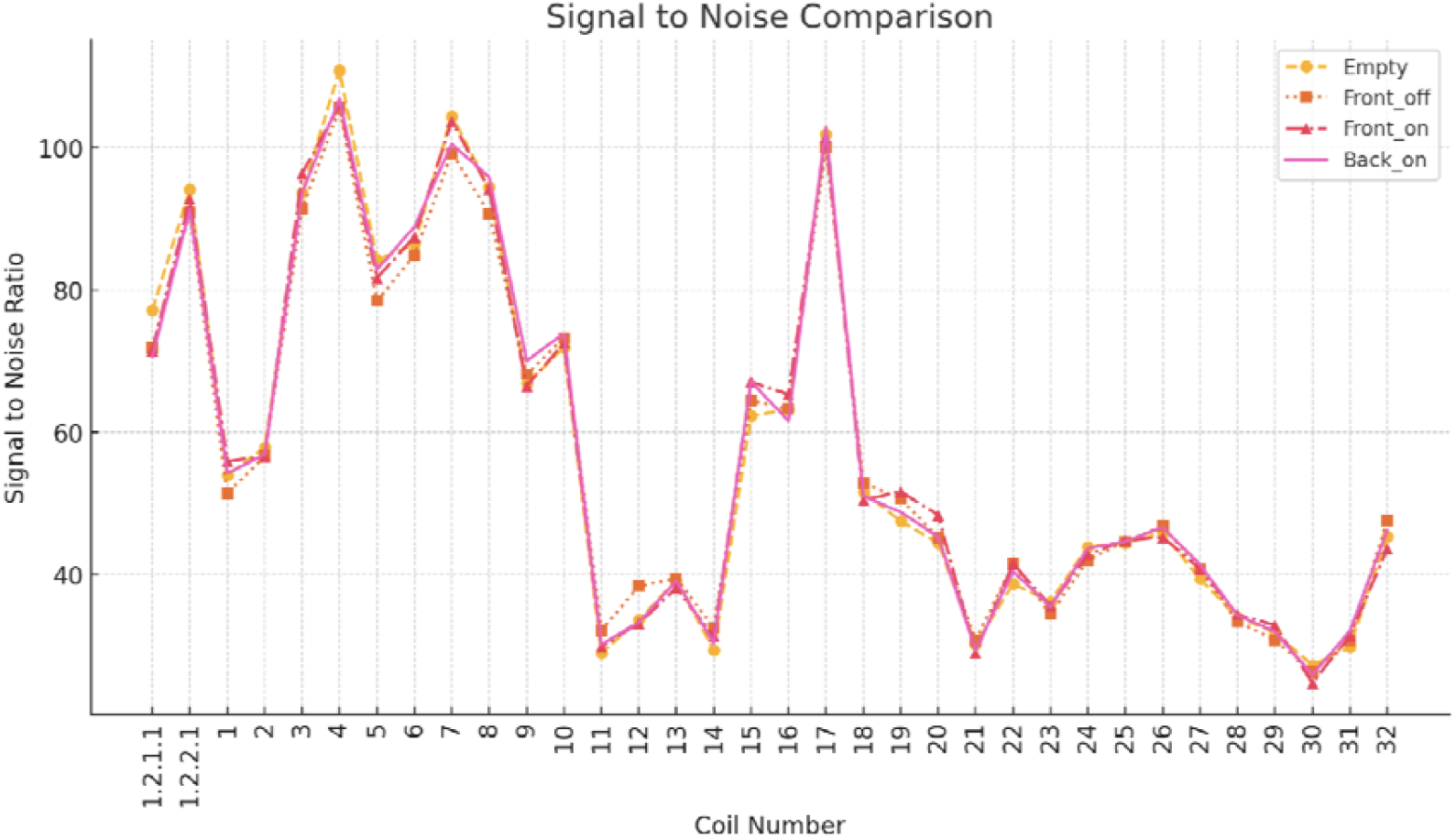
Quality Assurance of Head Phantom Signal to Noise Comparison conditions: no camera or stand (empty) camera off in front of room (front_off), camera on in front of the room (front_on), camera on in the back of the room (back_on).

## Notes

### Competing Interest Statement

The authors have declared no competing interest.

### Funding Statement

This study did not receive any funding

### Author Declarations

The Institutional Review Board of Brown University gave ethical approval for this work (protocol number 2262636-2)

